# The management of post-palatoplasty haemorrhage: A survey of cleft surgeons in Great Britain and Ireland and recommendations for local protocol implementation

**DOI:** 10.1101/2025.05.09.25327326

**Authors:** Bhavika Khera, Marc C. Swan, Guy Thorburn, Matthew Fell

## Abstract

**Objective:** To investigate strategies used by cleft surgeons to manage post-palatoplasty haemorrhage.

**Design:** Online cross-sectional survey

**Setting:** Great Britain and Ireland

**Participants:** 23 of 36 Cleft Surgeons (64% response rate)

**Results:** 16% of respondents indicated having a protocol for managing post-operative bleeds. There was an emphasis on the importance of preventative strategies to reduce the risk of post-operative bleeds occurring. Amongst the variety of non-surgical management options, systemic tranexamic acid and direct pressure were considered the most important. The tendency to prescribe antibiotics for secondary bleeds indicates a perceived difference in the nature or risk associated with these compared to primary bleeds. There was agreement that once identified, the management of post-operative bleeds should be proactive with a low threshold to return to theatre in the presence of an active intra-oral bleed.

**Conclusions:** We suggest a template with considerations that can be adapted into a local haemorrhage protocol for individual cleft units.

## Background

Post-operative haemorrhage following any form of intra-oral surgery is potentially life-threatening due to the risk of airway obstruction. This scenario can be particularly challenging in the paediatric setting, where the anatomy of the airway is smaller. The occurrence of post-operative intra-oral bleeds in the paediatric population has been well documented following tonsillectomy. Bleeding after tonsillectomy is commonly classified in the literature as primary haemorrhage if occurring less than 24 hours after surgery (usually caused by an uncontrolled bleeding vessel) and secondary haemorrhage if occurring more than 24 hours after surgery (may be due to inflammation or infection that has led to bleeding in traumatised tissue).^1^

In the cleft population, post-operative intra-oral bleeds can occur after both palatoplasty (including of primary cleft palate reconstruction, cleft lip reconstruction with vomer flap, buccinator flaps and secondary palate reconstruction) and pharyngoplasty (including sphincter-like pharyngoplasties and posterior pharyngeal flap).^2^ The occurrence of post-operative bleeding is fortunately thought to be low with a prevalence of 1-5% reported in the literature.^3–5^ Intra-operative strategies to minimise the risk of post-operative bleeding, including meticulous surgical technique and the use of haemostatic agents such as adrenaline infiltration and intravenous tranexamic acid have been shown to be effective.^6^

In the event of a post-operative bleed, it is important that the attending medical team can assess the patient and instigate effective management to achieve rapid cessation of the bleed. A clinical protocol (also known as a plan, pathway or guideline) is a tool to guide evidence-based healthcare.^7^ A protocol aims to standardise care and has the potential to streamline multidisciplinary clinical practice by detailing steps of management.^8^

Cleft care in Great Britain and Ireland has been centralised for the past 25 years in a ‘hub and spoke’ model with care provided by 13 cleft teams working within 17 surgical sites.^9^ Our aim was first to investigate strategies used by cleft surgeons to manage post-palatoplasty haemorrhage in Great Britain and Ireland and second to provide a recommended structure for the instigation of a local management protocol for the management of post-palatoplasty haemorrhage.

## Methods

### Survey design and distribution

An online survey was developed using Google Forms with reference to published guidance.^10^ To identify current practices of cleft surgeons in the scenario of a post-op bleed, questions were mapped to the three broad domains of (1) the existence of a bleeding protocol (2) interventions prior to theatre and (3) strategies if return to theatre was required. The final survey comprised 16 questions and included a mixture of single answer and free text options (see supplementary figure). The survey was circulated in electronic format in April 2024 via the Cleft Surgeons of Great Britain and Ireland online communication group, in which all current cleft surgeons and recently retired cleft surgeon are members.

### Presentation of survey results and further discussion

The results from the online survey were presented at the bi-annual Great Britain and Ireland cleft surgery clinical excellence network (CEN) meeting. Open discussion was documented. A post-palatoplasty haemorrhage template was designed and circulated around the CEN forum for feedback.

### Data Analysis

Survey responses were reported as proportions in line with the CHERRIES checklist for web-based surveys ^11^.

## Results

### Survey Demographics and Responses

The survey garnered responses from 25 participants, comprising 23 current cleft surgeons out of a total of 36 substantive cleft surgeons in Great Britain and Ireland, resulting in a 64% response rate. Additionally, one response each was received from a cleft surgery fellow and a retired cleft surgeon. The survey covered 12 of the 13 cleft services and 15 of the 17 surgical sites (Figure 1).

**Figure 1:**
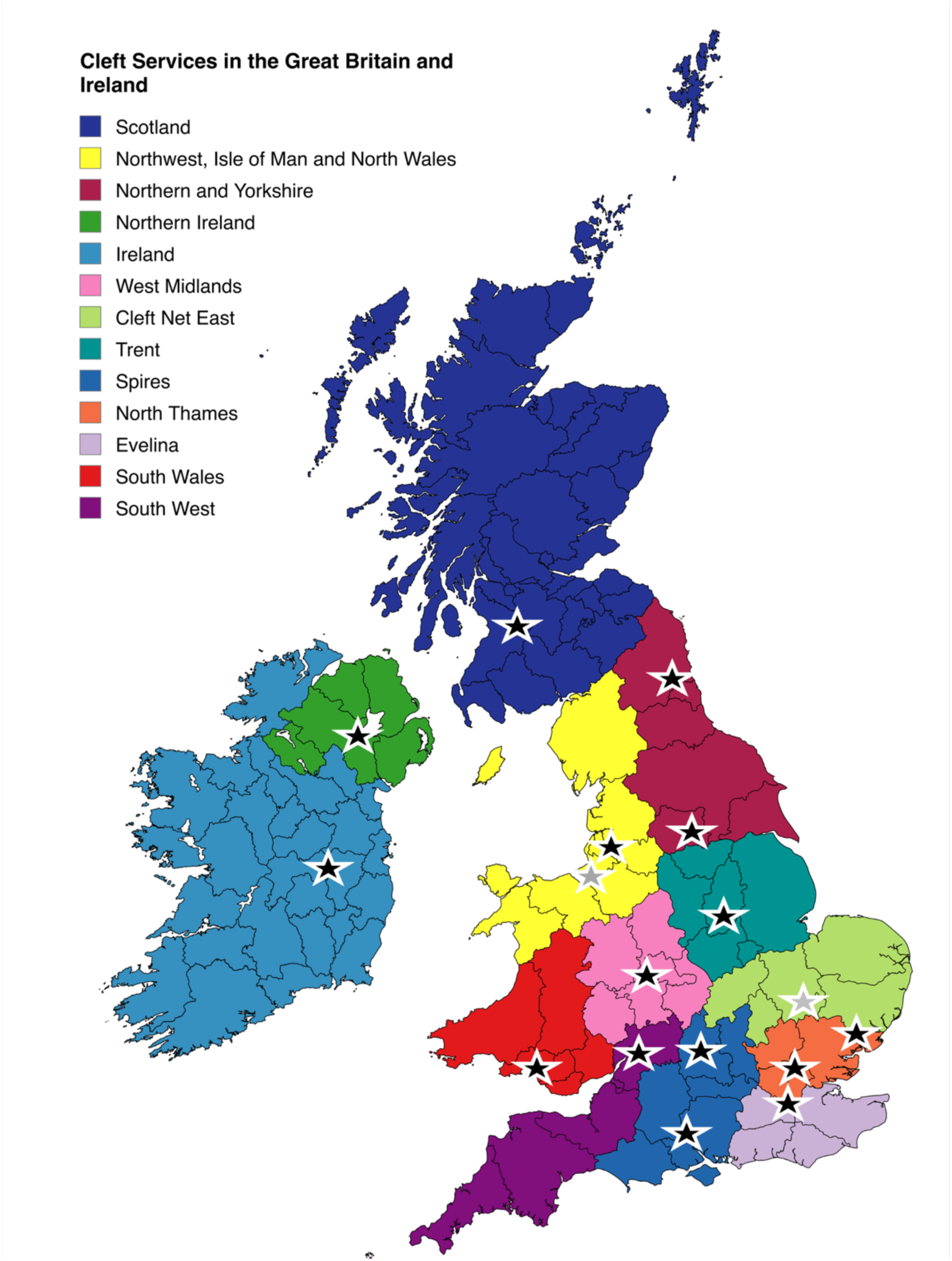
The 13 cleft services in Great Britain and Ireland. 17 Surgical sites are marked with stars. Black stars indicate the 15 surgical sites where responses were received and grey stars indicate the 2 services where responses were not received.

The on-call arrangement for post-operative cleft patients was the parent surgical specialty (Plastic or Maxillofacial Surgery) for 80% (20/25). A separate formalized cleft on-call rota was reported by 16% (4/25) whereas 4% (1/25) highlighted a lack of formalized on call cover, which was noted as a risk. 16% (4/25) of respondents indicated having a protocol for managing post-operative bleeds, spanning three separate surgical sites.

When asked to evaluate the importance of non-surgical interventions for managing post-operative bleeds, systemic tranexamic acid (TXA) and direct pressure emerged as the two most important interventions among the five options provided (Systemic TXA, topical TXA, nebulised adrenaline, topical adrenaline and direct pressure as shown in Figure 2). The survey explored differing approaches to primary versus secondary bleeds, revealing a split in responses: half of the respondents saw no difference in approach, while the remainder suggested that primary bleeds would more likely necessitate a return to theatre with comments indicating a higher likelihood to prescribe antibiotics for secondary bleeds. There was a clear consensus that a significant post-operative bleed at any timepoint would necessitate a prompt return to theatre. Regarding the use of packing or Surgicel in awake paediatric patients, 20% (5/25) of respondents would consider it, often depending on the patient’s age and cooperation, while 80% (20/25) would not. Furthermore, 88% (22/25) of respondents were comfortable with healthcare professionals suctioning the oral cavity, whereas 12% (3/25) preferred to avoid suctioning on the ward.

**Figure 2:**
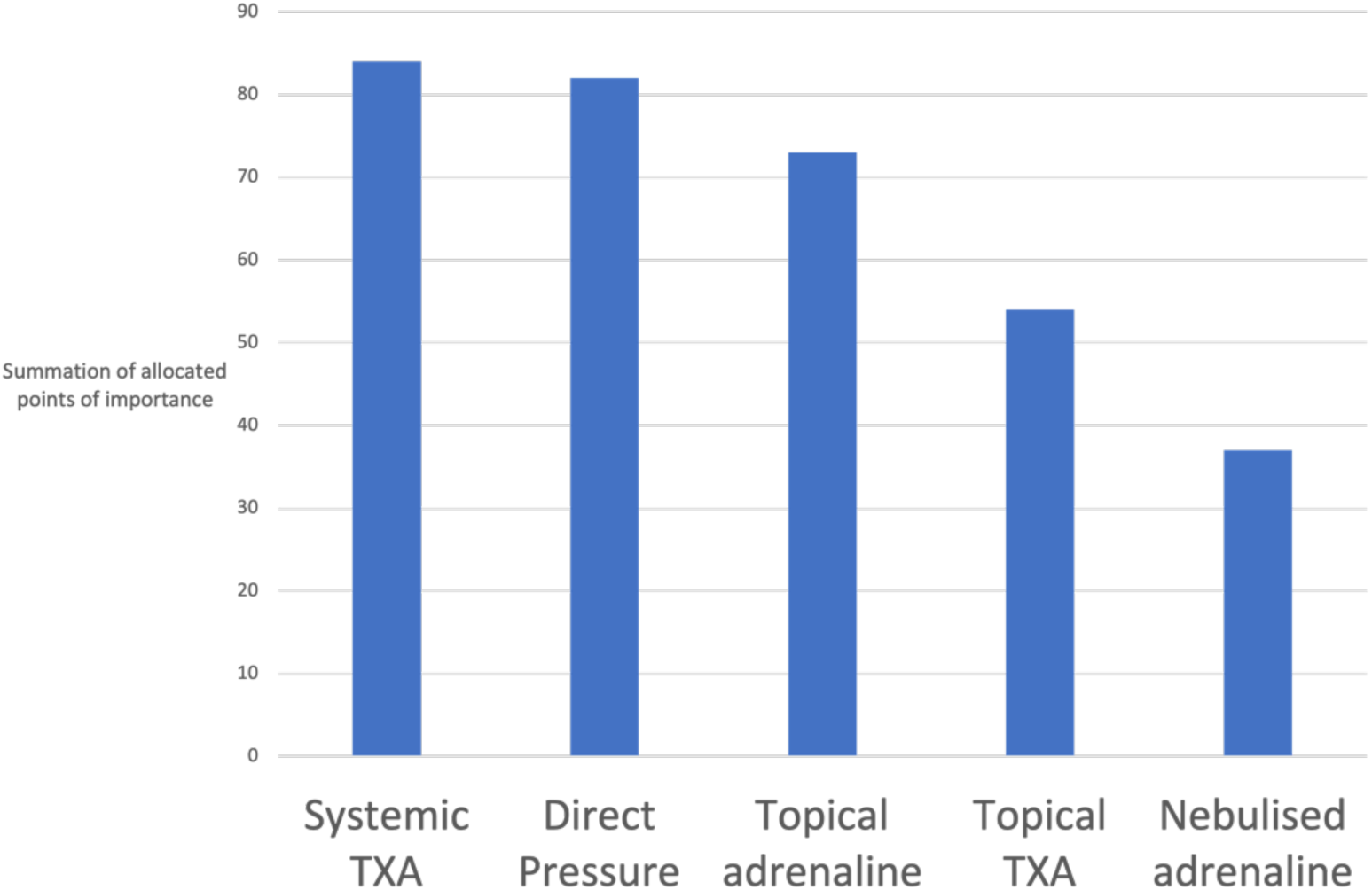
The importance of non-operative interventions for the management of post-op bleeds. Summation of participant scores of 1(least useful) to 5 (most useful) for each of the 5 intervention options

The importance of meticulous haemostasis was emphasized in white-space responses to prevent post-operative bleeds, with a low threshold for returning to theatre when a bleed was recognized. In situations where a return to theatre was indicated, respondents highlighted the necessity of involving an appropriately trained anaesthetist due to the potentially challenging nature of airway management in these cases. If a cleft surgeon was available, there was a strong consensus that they should manage the scenario, with any grade of assistance deemed acceptable. However, if a cleft surgeon was unavailable, responses varied regarding who should perform the procedure, likely influenced by existing on-call agreements. The most common response was to involve the on-call Plastic Surgeon in 36% (9/25), although concerns were raised about their comfort with inserting oral gags. Only 4% (1/25) advocated for a combined approach (i.e. on call Plastic, Maxillofacial and/or ENT surgeon), while 24% (6/25) believed the decision should depend on the expertise available.

### CEN meeting demographics and discussion

The discussion highlighted several key points regarding the management of post-palatoplasty haemorrhage. Centralised cleft care in Great Britain and Ireland is organised under a ‘hub and spoke’ model.^9^ It is plausible that patients may attend a ‘spoke’ peripheral hospital with a post-operative bleed where cleft surgeons are not present. The importance of collaboration and communication with ENT specialists in peripheral ‘spoke’ hospitals was emphasised, as they tend to be experienced at managing oral bleeds, particularly in the context of tonsillectomy, and are skilled in the use of mouth gags. Furthermore, ensuring that surgical trainees are familiar with essential skills including gag insertion, palate suturing, and effective bleeding control. The need for meticulous planning of operations was highlighted, particularly in anticipation of surgeon absences due to holidays, with recommendations to communicate these absences well in advance to ensure adequate on call cover and appropriate case selection. The discussion also stressed the importance of developing local protocols, cautioning against the use of generalised guidance documents as punitive measures if not suited to the local constraints.

Secondary bleeds were identified as potentially complex to both recognise and manage in a timely manner, necessitating a proactive approach. Pharyngoplasty procedures were noted as particularly challenging, due to the inaccessibility of the surgical site, underscoring the need for specialized training and preparation.

## Discussion

### Summary of main findings

In this cross-sectional survey of cleft surgeons in Great Britain and Ireland, we report differences of opinion surrounding specific aspects of managing post-operative bleeding, yet strong themes and areas of consensus emerged regarding management principles. There was an emphasis on the importance of preventative strategies, such as meticulous intra-operative haemostasis, to reduce the risk of post-operative bleeds occurring. There was agreement that once identified, the management of post-operative bleeds should be proactive with a low threshold to return to theatre in the presence of an active intra-oral bleed. The importance on training residents indicates a recognition of the need to equip surgical trainees with the essential skills for managing post-operative complications.

Amongst the variety of non-surgical management options, systemic TXA and direct pressure were considered the most important. There was divergence of opinion regarding recommending healthcare professionals to perform airway suction in the presence of a bleed and a reluctance to use packing in awake paediatric patients, likely due to concerns about patient comfort and cooperation. The tendency to prescribe antibiotics for secondary bleeds indicates a perceived difference in the nature or risk associated with these compared to primary bleeds. Secondary bleeds were considered a specific area of concern due to their propensity to occur when the patient was at home and potentially at considerable distance from the centralised cleft team ‘hub’.

### Strengths and limitations

The survey’s high response rate of 64% indicates a robust level of engagement among cleft surgeons, providing a credible snapshot of current practices across Great Britain and Ireland. Responses from 12 out of 13 cleft services suggest that the findings are broadly representative of regional practices. The survey’s scope on both surgical and non-surgical interventions offers a holistic view of management strategies, highlighting key areas of consensus while also identifying areas of variability and potential improvement.

Notable limitations include the reliance on self-reported data that may introduce response bias, as individuals with strong opinions or specific experiences may have been more likely to participate. The qualitative nature of some responses, particularly regarding the management of primary versus secondary bleeds, may lead to variability in interpretation and lack of quantitative precision. The survey reflects local variability of certain practices in Great Britain and Ireland, such as the choice of alternate surgeon when a cleft surgeon is unavailable, which may limit the applicability of the findings across wider clinical settings.

### Interpretation and recommendations

There is a paucity of data regarding post-palatoplasty haemorrhage, particular late secondary bleeds.^12^ Secondary bleeds pose unique challenges due to their potential occurrence at a distance from specialized cleft centres, often leading to initial presentation at peripheral hospitals. Whilst peripheral hospitals may lack specialised cleft services, they often have health professionals with airway training, such as ENT surgeons, which provides an opportunity for a network of coordinated emergency care to be delivered if a well communicated strategy has been devised.

The low proportion of cleft units with a post-operative bleeding protocol in place highlights a significant gap in standardized emergency preparedness and response strategies. The absence of formalized systems can lead to inconsistent management of post-operative complications, potentially affecting patient outcomes. This underscores the need for structured and reliable systems to ensure timely and effective responses to emergency situations. Whilst the variability in responses regarding the management of bleeds is perhaps not surprising amongst a group of highly specialised surgeons, this further highlights the necessity for clear guidance and training so that medical professionals on the front line can ensure consistent care.

Responders were clear that any guidance should be locally driven because without taking the local context into account, specific recommendations may be unfeasible under local conditions.^13^ The focus on local protocols suggests a desire to balance standardized guidelines with a flexibility to accommodate specific service needs, thus avoiding punitive measures for non-compliance.^14^

To address some of the challenges discussed following this survey, we have developed a template for the creation of a local guideline to manage post-palatoplasty haemorrhage (see Table 1). This acknowledges the impracticalities of creating a national protocol, and instead the proposed template highlights suggestions that can be adapted into a protocol for each unit depending on the resources available. The template covers three key areas:

1. How to recognise a post-operative bleed
2. Who to call for help
3. What management interventions to instigate using the well-recognised ABC (Airway/Breathing/Circulation) resuscitation format.

**Table 1:**
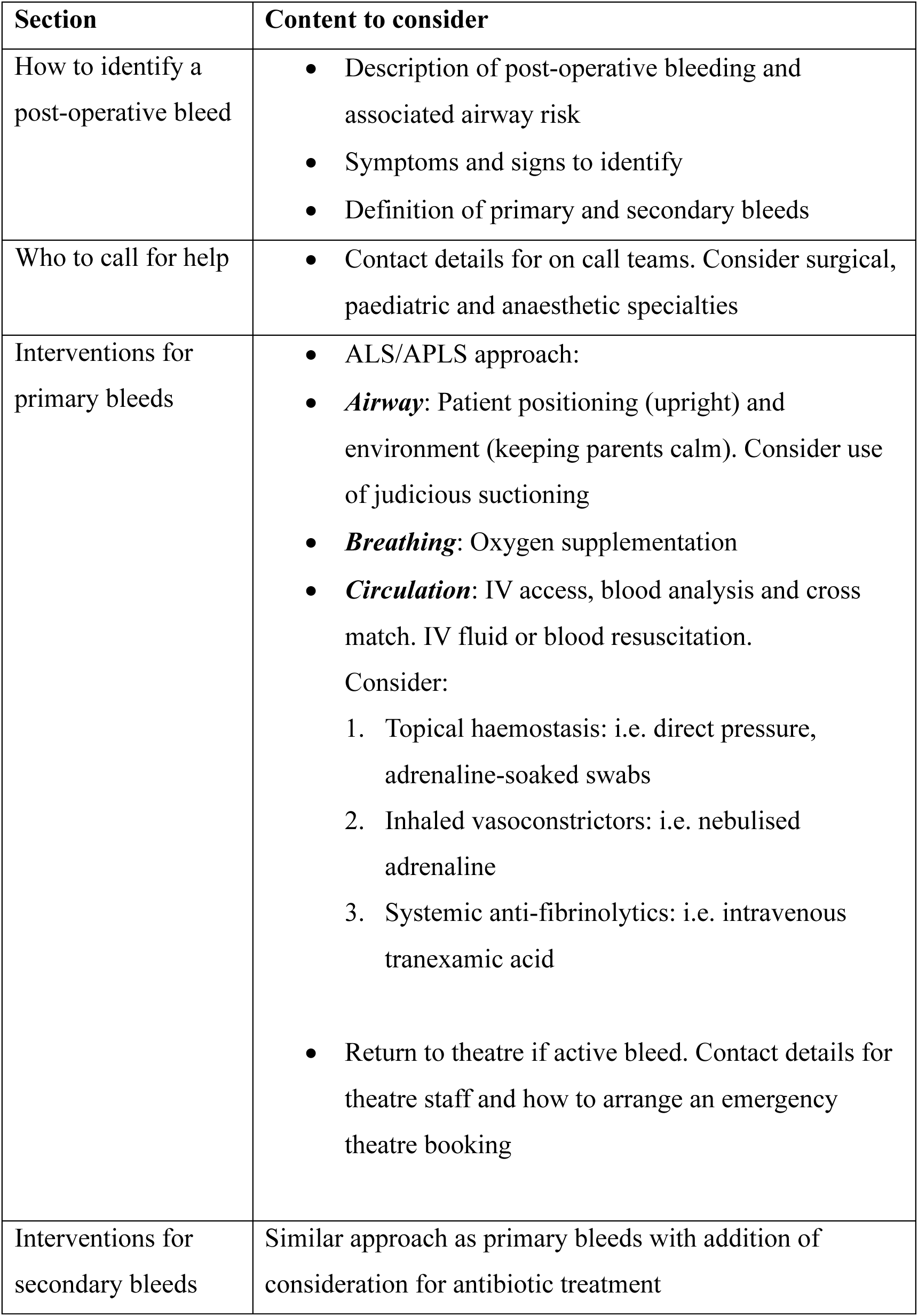
A template to assist with the creation of a local post-palatoplasty haemorrhage guideline.

Further recommendations to address the highlighted issues include targeted simulated training for healthcare professionals in both cleft centres and peripheral hospitals. This training should focus on the identification and management of post-palatoplasty haemorrhage, the challenges of late secondary bleeds and managing airway complications with essential interventions, such as gag insertion and haemostasis. Emergency response systems can enable rapid communication and coordination between peripheral healthcare centres and centralised cleft centres when a late secondary bleed is identified. Furthermore, implementing systems for collecting and analysing data on post-palatoplasty haemorrhage cases, particularly late secondary bleeds can help identify trends and areas for improvement. This data can be used to refine protocols and training programs, ensuring that they remain aligned with best practices and emerging evidence.

### Conclusions

This survey highlighted expected variation in local practices and surgeons’ preference regarding the management of post-palatoplasty haemorrhage in Great Britain and Ireland. The utility of a well-planned and coordinated approach in the event of a post-operative bleed is clearly evident. We suggest a template with considerations that be adapted into a local haemorrhage protocol for individual cleft units.

## Data Availability

All data produced in the present study are available upon reasonable request to the authors

## Acknowledgements

Cleft Surgery Clinical Excellence Network (CEN) Group of Great Britain and Ireland

## Supplementary Material

Supplementary Figure 1: The online survey

## Cleft Postoperative Bleeding Survey

Thank you for completing this survey. We would like to know about your experience and any protocols for the management of postoperative bleeding after cleft surgery (palate repair and pharyngoplasty).

1. Email * ______________________
2. What is your professional status? *Mark only one oval.* 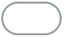 Cleft Surgeon 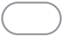 Retired Cleft Surgeon 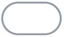 Training Cleft Surgeon (Fellow or registrar) 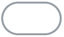 Other:_____________
3. Which Cleft Unit do you work in? *Mark only one oval.* 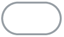 South Wales 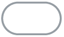 South West 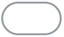 Spires 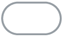 South Thames 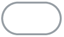 North Thames 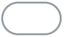 Cleft Net East 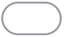 Trent 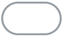 West Midlands 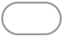 North West Manchester 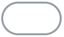 North West Liverpool 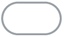 Northern Region Newcastle 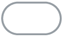 Northern Region Leeds 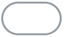 Scotland 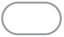 Belfast 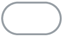 Dublin 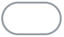 Other:_____________
4. Have you had personal experience of managing a patient with post-operative bleeding following palatal or pharyngeal surgery? *Mark only one oval.* 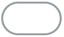 Yes > 5 patients 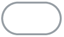 Yes 3-5 patients 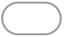 Yes < 2 patients 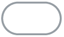 No
5. Does your cleft unit have a written protocol for the management of post-operative bleeding following palatal or pharyngeal surgery? *Mark only one oval.* 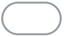 Yes 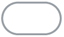 No
6. What is the oncall set up for out-of-hours cleft emergencies in your unit? *Mark only one oval.* 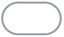 Cleft surgeons have their own on-call rota (ie formal job-planned rota) 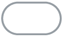 Plastic Surgery provide cover 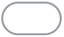 Maxillofacial Surgery provide cover 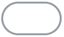 ENT provide cover 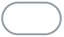 Combination of specialty surgical teams provide cover 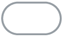 Other:_____________
7. When considering post-op bleeds, do you have a different management approach for primary bleeds (<24 hours post-op) and secondary bleeds (>24 hours post-op)? *Mark only one oval.* 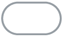 Yes: more likely to return to theatre for primary bleeds 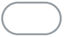 Yes: for other reasons - please specify below 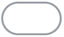 No
8. Please specify differences in approach to primary and secondary bleeds _________________________ **Interventions prior to theatre** For each intervention, please select how important you find them to be
9. Systemic tranexamic acid *Mark only one oval.*

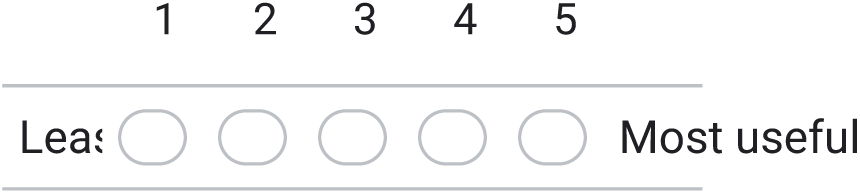
10. Topical tranexamic acid *Mark only one oval.*

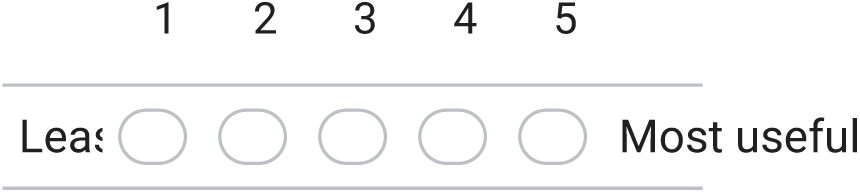
11. Nebulised adrenaline *Mark only one oval.*

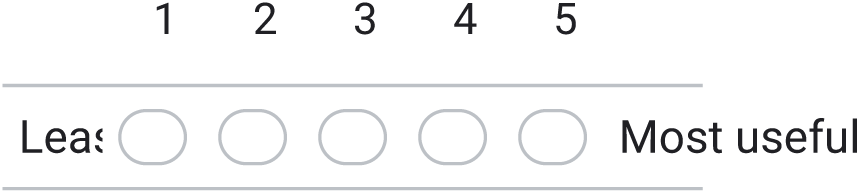
12. Topical adrenaline *Mark only one oval.*

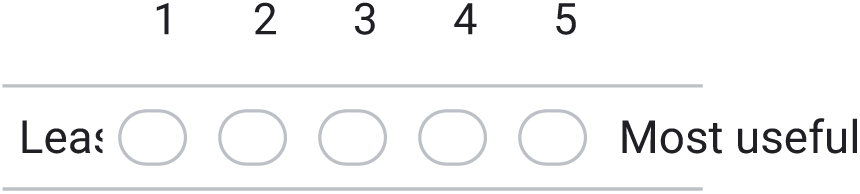
13. Direct pressure *Mark only one oval.*

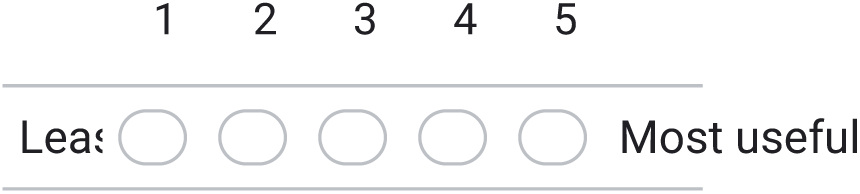
14. Would you consider packing or Surgicel in an awake paediatric patient with a postoperative bleed? *Mark only one oval.* 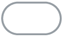 Yes 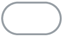 No 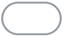 Other:_____________
15. Would you permit another healthcare professional to suction the oral cavity prior to your arrival? *Mark only one oval.* 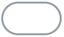 Yes 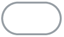 No **Return to theatre** If there is an indication for a return to theatre, please consider the following scenarios where a cleft surgeon **IS** or **IS NOT** available
16. If a cleft surgeon **IS** available for theatre, who else would you want available to assist? *Mark only one oval.* 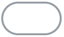 Specialty surgical trainee or fellow 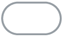 Oncall Consultant Maxillofacial Surgeon 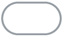 Oncall Consultant Plastic Surgeon 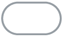 Oncall Consultant ENT Surgeon 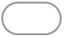 Other:_____________
17. If a cleft surgeon **IS NOT** available, who would you want doing the case? *Mark only one oval.* 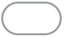 Oncall ENT Surgeon 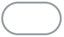 Oncall Plastic Surgeon 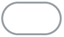 Oncall Maxillofacial Surgeon 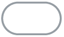 Combined approach with at least 2 consultant surgeons 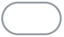 Other:_____________
18. Do you have any other tips or thoughts about protocols for the management of post-operative bleeds in palatal or pharyngeal cleft surgery? ___________________________________ ___________________________________ ___________________________________ ___________________________________ ___________________________________

